# Contact in context: Animal profiles, human activities, and land use histories shape human-animal contacts with implications for zoonotic spillover in the Democratic Republic of Congo

**DOI:** 10.64898/2025.12.07.25341615

**Authors:** Romain Duda, Victor Narat, Thibaut Langlois, Robert Monfura, Marc Allassonnière Tang, Placide Mbala, Claude Monghiemo, Tamara Giles-Vernick

## Abstract

Pathogenic spillovers from animals into humans have catalyzed epidemics throughout history. They result from multiple factors. One such factor, human-animal contact, remains poorly understood. Current studies often neglect variability in human-animal engagements across ecological zones and the broader processes bringing people, animals, and pathogens into engagement. We investigated factors and longer-term processes shaping human-animal contacts and risks of zoonotic spillover in a region experiencing landscape fragmentation.

We conducted our mixed-methods investigation in three villages along an ecological gradient of forest fragmentation in the Democratic Republic of Congo (DRC), a hotspot of biodiversity and disease emergence. Among 24 village participants, we collected daily activities and contacts with highly diverse animal species to evaluate the types and frequencies of these contacts. We developed a cluster analysis to categorize classes of animals according to type and frequency of contact. We also conducted transects to estimate animal species abundance according to village proximity. We tested the influence of animal species abundance, human activities, gender, and village on human-animal contact frequency. We conducted ethnographic and ethnohistorical interviews and observations to explore changing human-animal relations.

Participants had physical and environmental contact with 61 different animal species. We found three classes of animal species with which participants had most frequent physical and environmental contact. Historical processes and human activities, avoidance toward some animal species, and relative abundance of species contribute to shape contemporary human-animal contacts, and more broadly, potential risks of exposures to zoonotic pathogens. Our modeling of gender, village, relative abundance and human activities on animals clustered by contact frequency, however, yielded few predictors of contact frequency.

We identify factors and processes associated with human-animal contacts in an ecologically varied zone and its categorization of contact profiles. Future studies should explore a wider array of human-animal contacts and situate them in their historical contexts.

**Author Summary:** Pathogenic spillovers from animals into humans have catalyzed epidemics throughout history. These spillovers result from many factors, although one –– human-animal contact –– is poorly understood. The variability of human-animal interactions and historical changes shaping interactions between people, animals, and pathogens are not well addressed. Our mixed-methods study explored factors and longer-term processes affecting human-animal contacts and risks of spillover in a fragmented forest of the Democratic Republic of Congo.

We found that participants had physical and environmental contact with 61 different animal species. We identified three classes of animal species with which participants had most frequent physical and environmental contact. These classes were shaped by three major factors: historical changes affecting human activities and ecologies; human preferences to avoid certain species; and relative abundance of animal species. More broadly, these classes reflected potential risks of exposures to zoonotic pathogens. Although our model to predict how gender, village, relative abundance and human activities influenced these animal classes clustered by contact frequency, it yielded few predictors. Our study did, however, identify factors and processes associated with human-animal contacts in an fragmented forest zone. We recommend that future studies explore a wider array of human-animal contacts, situating them in their historical contexts.

## Introduction

Pathogenic spillovers from animals into human populations (anthropozoonoses) have long constituted a threat to public health, resulting in the transmission of several neglected tropical diseases (NTDs) and in some cases, catalyzing epidemics and pandemics [1,2]. Recent mpox and COVID-19 pandemics, for instance, have had their beginnings in nonhuman animals, but their pathways into human populations are complex, shaped by multiple factors, including phylogenetic proximity between hosts, a pathogen’s adaptive capacity, and animal ecologies and behaviors that may intersect with human practices to shape cross-species contacts [3]. Anthropogenic transformations, from climatic and resulting ecological changes, to habitat fragmentation, demographic expansion, hunting, butchering and wildlife trade, all shape human-animal contact [4–6]. Human-animal contact, although a critical factor in facilitating zoonotic spillovers, is still poorly understood at a fine scale. Prior research has tended to focus on large scale changes, from intensive agricultural production to commercialized animal production to deforestation, that occasion biodiversity loss and modify patterns of human-animal contact [7–9]. Nevertheless, zoonotic spillovers are not only the consequence of broad scale changes, but also the result of specific human-animal-pathogen interactions at specific sites and moments [10,11].

These specificities, however, have not been fully tackled in literatures on human-animal contact. Certain studies have not defined or used “contact” consistently [12]. Large-scale explanations for and mechanisms of spillover, moreover, do not account for granular human-animal interactions; they thus neglect why zones experiencing anthropogenic disturbance do not all produce traceable zoonotic spillovers. Granular, small-scale investigations shed light on a range of reported human-animal contacts at times a limited group of animals (nonhuman primates); a few have productively explored human contacts with a wide range of pre-determined animal species to address the complex pathways that pathogens may follow in host spillovers [13–15]. To our knowledge, just one recent study has evaluated contact frequency and integrated quantitative and qualitative data collection tools to elucidate the types and frequencies of human-human and human-animal contacts [16]. Most studies, however, still neglect granular insight into the rich contexts of these contact events, the variability of human-animal engagements, and the broader historical, socio-cultural, economic, and ecological processes and practices bringing people, animals, and pathogens into engagement.

Generating deeper insight into human risks of zoonotic transmission necessitates that we expand our scope and methods to undertake a socio-ecological investigation. More broadly, complex relations among undomesticated and domesticated animals, human societies, and pathogens can shape zoonotic transmission pathways [17–19]. Interspecies contact can result from relations of predation, competition, mutualism, or avoidance, from overlapping ecological niches and sharing of resources and habitat, as well as coincidental nocturnal or diurnal activities. Species diversity and abundance can further influence contacts and indirectly pathogenic transmission [20]; large-scale climatic and ecological transformations may intersect with smaller scale ecological changes, which in turn can shape species distribution, density, and behavior, thus affecting interspecies contacts [21,22].

This manuscript therefore asks what factors, conditions, and longer-term processes have shaped human-animal contacts and thus risks of zoonotic spillover in a fragmented landscape? To tackle this question, we conducted a study on the Congo basin forest edge of the Democratic Republic of Congo (DRC), a hotspot of biodiversity and disease emergence. Our mixed-methods investigation brought together ecological and anthropological tools to explore human-animal contacts at a local scale. Our study analyzed quantitatively and qualitatively the diversity of domesticated, commensal, and undomesticated animal species with which human inhabitants had contact, as well as types and frequencies of these human-animal contacts. We examined the influence of specific animal relative densities on contact types and frequencies and situated this contact analysis in broader historical changes in land use, hunting and trapping, and animal production. We also explored comparatively whether animal abundance, human activities, village sites, or gender affected contact frequencies. This article thus offers a broad analysis of contact and the historical processes shaping it, to generate a comprehensive depiction of human exposures to animals in a fragmented habitat in Central Africa.

## Material and Methods

### Study sites and study period

We conducted field research from March to August 2019 and in September 2021 in three villages (here anonymized as villages A, B and C) located in Northern Bateke chiefdom (Chefferie des Bateke Nord) in the Bolobo Territory, Maï-Ndombe Province, Democratic Republic of the Congo (DRC) (Fig 1). Situated along the southwestern edge of the Congo basin forest, this area is characterized by a fragmented forest-savanna mosaic, composed of lowland rainforest and herbaceous and woodland savannas [23]. Characteristics of the three villages are summarized in Table 1.

**Fig 1:**
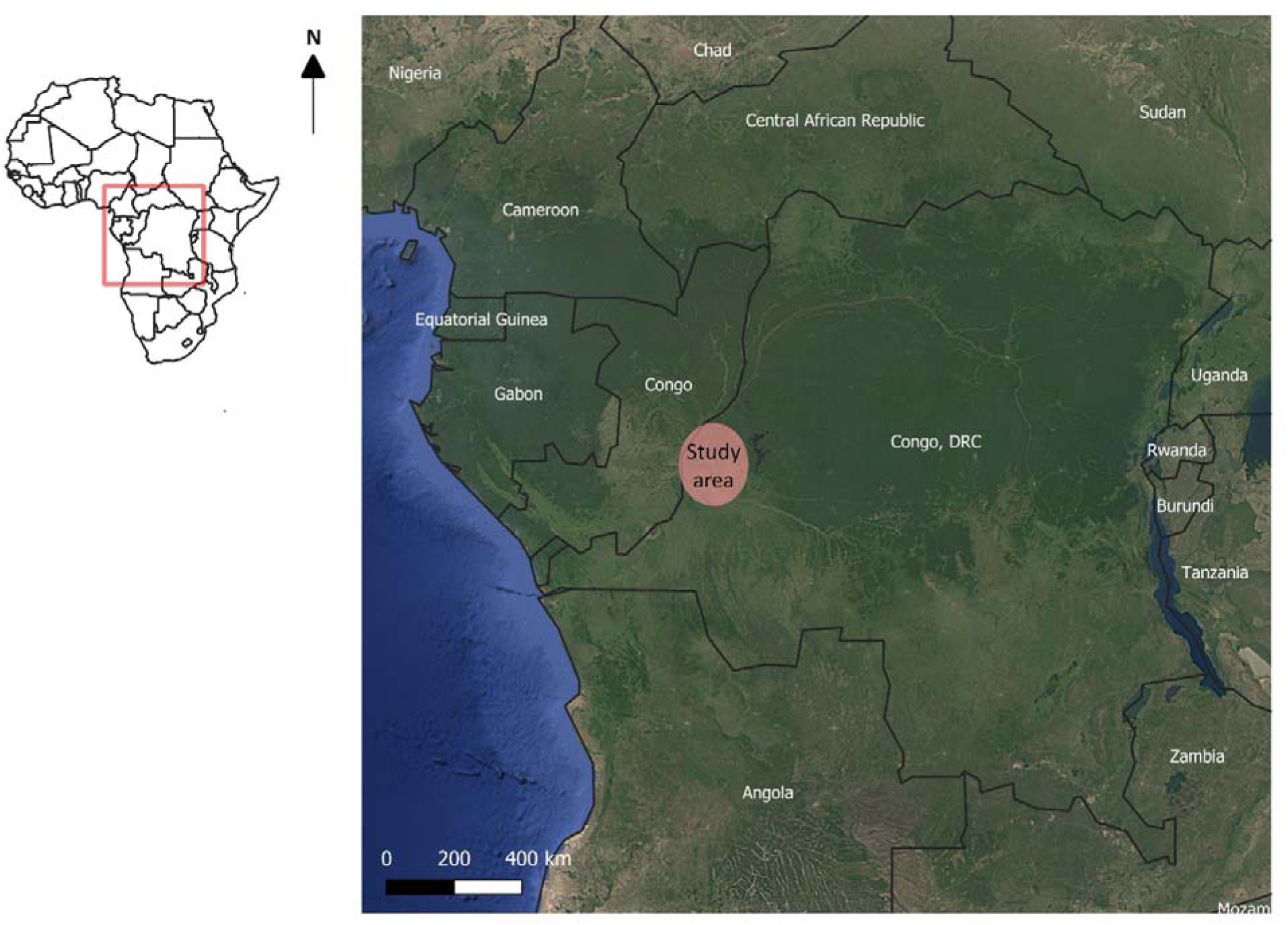
Map of the study area. Base map Google Earth®.

**Table 1:**
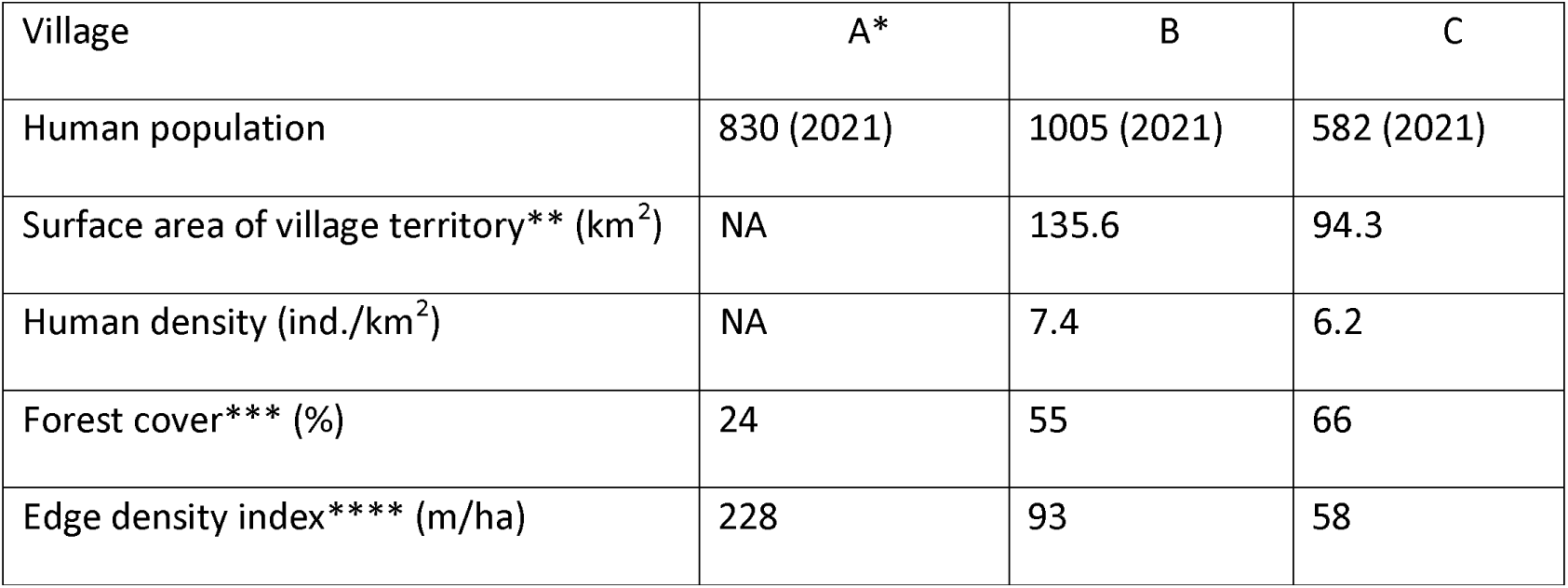
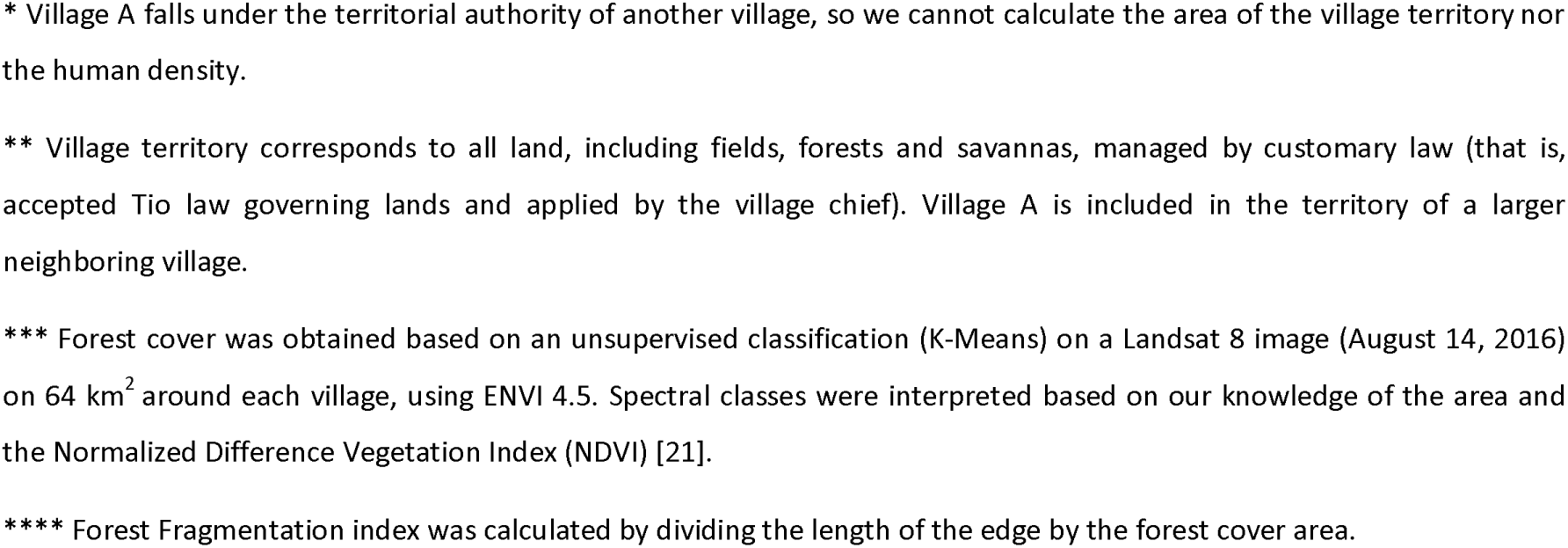
Characteristics of the three villages targeted in this study.

| Village | A* | B | C |
| --- | --- | --- | --- |
| Human population | 830 (2021) | 1005 (2021) | 582 (2021) |
| Surface area of village territory** (km<sup>2</sup>) | NA | 135.6 | 94.3 |
| Human density (ind./km<sup>2</sup>) | NA | 7.4 | 6.2 |
| Forest cover*** (%) | 24 | 55 | 66 |
| Edge density index**** (m/ha) | 228 | 93 | 58 |

\* Village A falls under the territorial authority of another village, so we cannot calculate the area of the village territory nor the human density.
\*\* Village territory corresponds to all land, including fields, forests and savannas, managed by customary law (that is, accepted Tio law governing lands and applied by the village chief). Village A is included in the territory of a larger neighboring village.
\*\*\* Forest cover was obtained based on an unsupervised classification (K-Means) on a Landsat 8 image (August 14, 2016) on 64 km<sup>2</sup> around each village, using ENVI 4.5. Spectral classes were interpreted based on our knowledge of the area and the Normalized Difference Vegetation Index (NDVI) [21].
\*\*\*\* Forest Fragmentation index was calculated by dividing the length of the edge by the forest cover area.

The region’s mean annual pluviometry is 1625 mm, with a rainy season lasting from October to May and a dry season from June to September [24]. Its forests and savannas host highly diverse terrestrial and arboreal mammals such as antelopes, boars, rodents, carnivores, and nonhuman primates (including bonobos, *Pan paniscus*) [25]. The village also houses several domesticated species, notably chickens, goats, sheep, dogs and cats, as well as commensal species such as mice and bats. Extensive cattle farming occurs in the savanna zones.

Human population density is low (< 10 ind./km^2^), and human settlements are situated in savanna zones. Although one town on the banks of the Congo River has approximately 10,000 inhabitants, most village populations range between 250 and 3,000 people. The predominant ethnic group is Tio, part of the larger Teke (or Bateke) linguistic group which migrated from what is present-day Republic of Congo to this zone, likely during the 18^th^ century as a consequence of slave and other trades [26]. This population generally practices subsistence swidden agriculture, gathering, hunting, fishing, and engages in local trade [27].

For the past 25 years, this study area has also been the site of a community-based conservation project developed by the Congolese nongovernmental organization (NGO) Mbou-Mon-Tour [28]. Led through local, bottom-up governance, this project supports bonobo conservation, with three sites of habituated bonobos for eco-tourism and scientific research [28].

### Data collection

#### Animal contact and forest activity frequencies through a participatory approach

To evaluate the diverse types and frequencies of contact between humans and animals in near real-time, we adapted a participatory data collection tool previously developed by research team members [15]. Because participation was remunerated and thus highly sought after, we requested each village chief in our three sites to select ten participants (hereafter “volunteers”) per village (total 30), so that we avoided creating local tensions. Village chiefs thus chose participants from each site to convene to our inclusion criteria (adult >18 years old, healthy, resident of study village, regular practitioner of activities outside of the village). We encouraged chiefs to recruit individuals from different lineages and villages neighborhoods and to respect gender parity. Despite our guidelines, village chiefs (male authorities in a patriarchal society) selected more men than women. Men also generally had higher educational levels and more free time than women, and thus were more able to participate, even when we provided scribes for women who could not read or write. Additionally, more men than women volunteered to participate. Overall, our male participants outnumbered female participants (19 and 11, respectively).

All trained volunteers documented their animal contacts for five consecutive months, filling out two datasheets daily in their local language (Etio or Kiteke). The first datasheet documented any contact with an animal, either domesticated or not (excluding non-domestic birds, arthropods and fish); participants noted the species encountered or signs of its presence. Volunteers also documented the type(s) of contact taking place: environmental (participant saw feces, food remains, nest/hole/burrow and/or footprints), direct (participant saw animal alive or dead, or heard animal); or physical contact (participant hunted, transported, butchered, prepared/cooked, ate, purchased, or sold the animal). For the second data sheet, volunteers documented the activities performed that day (hunting, fishing, agriculture, gathering, cattle breeding, or no activity in the forest) (Supplementary Information 1a).

Data quality was ensured through multiple measures. Volunteers underwent training by Author 1, who monitored their study activities during the first and last months of data collection, as well as a full-time research assistant (Author 4), who supervised volunteer data collection during the rest of the participatory study. Because several volunteers could not write, we trained a young literate family member to assist them in filling out the data sheets. We also regularly verified information collected, requesting that volunteers orally recount their contacts with animals over the past 48 hours. Of the 30 volunteers recruited, we excluded six because their data were unreliable or inconsistent. The final dataset included 24 volunteers (7 women, 17 men), with a mean age of 48 years old (+/− SD=18, range=22-80). We obtained a mean of 160 days with data on animal contacts per volunteer (+/− SD=5.3; range= 144-167).

#### Transect survey

We developed faunal transects for each village site to estimate the relative abundance of certain animal species for which we could obtain signs of presence. Developing an 8*8 km square, with each village at the square’s center, we then drew 2*2 km squares inside of the larger square, so that each resulting diagonal of the sub-squares constituted a transect. We then divided each transect into a 50-meter section. For each section, local trackers from the NGO Mbou-Mon-Tour were trained by Author 2 to record all animal traces. These trackers all had deep experience recognizing animal signs of presence. Certain sections of transects were not possible to access because landowners did not permit data collection. We therefore censused a total distance of 44.45 km, 41.50 km and 44.90 km for villages A, B and C respectively. Due to a data collection error in Village B, non-human primates were not registered there and thus were not included in the transect analyses.

#### Household survey for commensal species

Scientific studies show that certain housing infrastructures and building materials are associated with a higher frequency of synanthropic species (notably mice and bats) [29–31]. We therefore conducted a short questionnaire (Supplementary Information 1b) to evaluate the presence or absence of two major commensal species (*Mastomys natalensis* and *Pipistrella* sp.) inside houses, and to document housing types, namely the type of roof (straw or sheet metal) and walls (brick or mud), and the effects of the type of construction on commensal distribution. Over two days in each village, a research assistant conducted the survey with all household heads. We conducted surveys among 272 households total (99 in village A, 136 in village B, and 37 in village C).

#### Semi-structured interviews, participatory and direct observation

To gain greater insight into current human-animal contacts and the underlying historical dynamics shaping them, we collected anthropological data with village inhabitants using semi-structured interviews, informal interviews, focus group discussions, and participant-observation. Each interview and focus group lasted 30 to 60 minutes, with discussions focusing on local histories of hunting and wildlife dynamics and behavior, relations with commensal species, goat and sheep raising, poultry farming, dogs as hunters and guards, cattle raising, diet and food avoidances, habitat and landscape dynamics, and illnesses (SI 1c for semi-structured interview guides). We employed focus group discussions to validate vernacular species names (using [32]) and to understand local categorizations of animal species and ecological habitats. Participant-observations documented daily activities, including hunting, fishing, and gathering. We conducted 88 semi-structured interviews, 8 focus group discussions (average 5 persons/group), and around 30 hours of participant-observation.

For qualitative data collection, we recruited adult participants based on their willingness to participate in interviews, focus group discussions, and participant-observation. Our recruitment relied on snowball sampling; we asked our participants to identify other knowledgeable people who might be willing to participate. We followed local norms in prioritizing certain gender and age groups to find individuals with deep knowledge about specific subjects. Hence, for interviews about domestic animals, we sought out middle-aged and older men to discuss cattle-, goat-, and sheepherding, and women of all ages for chicken and duck raising. Interviews focusing on forest and savanna species were conducted with active or retired hunters (mostly men). For historical interviews, we sought out elders (mostly men, who are purveyors of such knowledge), born in one of the three villages and coming from different families. We conducted interviews to saturation, that is, until we gathered no new knowledge. We led all interviews and focus group discussions in French, which our research assistant translated directly into Etio. We recorded these exchanges if the participant consented, and for the few who refused, we took detailed notes. All recordings were transcribed and then translated into French by a native Etio speaker.

### Data analysis

#### Animal contact frequencies and clustering analysis

We developed our animal contact frequency and clustering analysis to provide broad insight into human exposures to animals, not a detailed examination of contact types and frequencies with specific species as a function of gender or village site. To that end, we developed an index of presence/absence per day to quantify the frequency of one type of contact for one animal species. We defined a “contact event” as one volunteer having one type of contact with one animal species on one date. Thus, if on a specific day a volunteer had three types of contact with one species, we would count these engagements as three distinct contact events. We calculated for each volunteers a frequency for all interactions “animal species*type of contact” with the following formula:

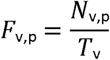

where *F*_v,p_ is the frequency of the parameter *p* (an interaction “animal species*type of contact”) for the volunteer *v*, *N*_v,p_ is number of days with a recording of the parameter *p* by the volunteer *v*, and *T*_v_ is the total number of days with data for the volunteer *v*.

The same method was used to calculate the frequency of each human activity, including different techniques used for hunting and fishing. We used the following formula:

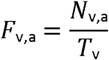

in which *F*_v,a_ is the frequency for the activity *a* for the volunteer *v*, *N*_v,a_ is number of days with a recording of the activity *a* by the volunteer *v*, and *T*_v_ is the total number of days with data for the volunteer *v*. Thus, for each volunteer, we obtained a frequency for each category of activity (hunting, fishing, agriculture, gathering, cattle raising) and a frequency for each specific technique of hunting and fishing.

To avoid imposing our own assumptions about animal species categories (e.g. “domestic” or “wild”), we used a cluster analysis to identify animal categories of contact profiles based on the contact frequency for each contact type. First, we performed a Principal Component Analysis (PCA) on mean contact frequencies for each type of contact and each animal species, so that we could visualize the correlations between contact types. We then reassigned the coordinates of the first two axes to each animal species and performed on these coordinates a hierarchical cluster analysis (HCA, R package Cluster [33]) with the Ward method [34]. To characterize each cluster obtained, we compared mean frequencies for the 14 contact types with the mean relative abundance between the three clusters, using a Kruskal Wallis test and a pairwise Wilcoxon test.

#### Influence of relative abundance index, gender and village on animal contact frequencies

We assumed that the closer to a village an animal species is, the higher the probability of contact with this species. On that assumption, we calculated a relative abundance index for each animal species, adapted from Narat et al. 2018 [15], with the following formula:

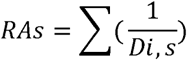

where *RAs* is the relative abundance for the species *s*, and *Di*, *s* is the Distance (km) of the sign of presence *i* for the species *s* recorded during transect survey.

We then built Generalized Linear Mixed Models (GLMMs) to test the influence of village, gender, relative abundance index, and frequencies of forest activities on contact frequency. We built one model for each interaction between the contact category obtained from the PCA and the clusters of animal species from the HCA. The response variable was the contact frequency, and the predictive variables (fixed effect) were village, gender, human activity frequency, and relative abundance of animal species. We controlled for volunteers and animal species as random effects. For each model, we compared the full model vs the null model: we compared whether a full model with fixed effects plus random effects yielded better results than a null model with only random effects. If the former could not explain more variance than the latter, it indicated that the predictive variables did not explain much of the variance of contact frequency. The details of the models and their interpretative plots are provided in the supplementary materials (Supplementary Information 2).

#### Commensal species and house type

We analyzed the presence of mice and bats in house and relations with housing materials. We conducted the statistical analysis by employing Fisher’s exact test for each variable individually, followed by a combination of variable tests.

#### Qualitative data on human-animal interactions

We imported transcripts and observation notes into qualitative data analysis software to code and label inductively text segments. We inductively coded by reading transcripts and notes closely and highlighting recurrent phrases related to human-animal interactions, developed codes to describe these texts (such as avoidance of animals; interactions with rodents; interactions inside house), and then refined these codes. Aggregating our codes, we identified themes that characterized the full qualitative dataset, such as contact through hunting; sharing norms; livestock farming; declining undomesticated animals. To enrich our interpretations and develop comprehensive insights, we triangulated results from our quantitative and qualitative approaches. We therefore present results by integrating our insights from both data types.

#### Software used for analysis

All statistical analyses were performed with R version 4.0.3 [35]. We used ade4, cluster and lmerTest packages to perform the PCA, HCA and GLMMs [33,36,37]. Spatial analyses determining the distance from village site of each sign of animal presence were performed with QGIS Madeira (version 3.4.3)[38]. We conducted our qualitative coding with NVivo 11 software (*Lumivero*, version 1.7.1 (1534)) [39].

### Ethics and research authorization

The research protocol, information notice, and consent forms were evaluated by and received ethical approvals from the Institut Pasteur Institutional Review Board (Decision No. IRB2018-08) and the National Committee of Health Ethics (Comité National d’Éthique de la Santé, CNES) of DRC (Decision no. ESP/CE/2019 and ESP/CE/045/2021). All potential participants received written and oral descriptions of the study, procedures for participation, and participant rights. All participants provided written informed consent. This study is registered on clinical trials database (n°NCT04012164). Before beginning our study, we presented all study aims and methods to village chiefs for their authorization to conduct our research.

## Results

Below we present a description of participants, current human activities affecting the array of contacts with different animal species, volunteer contacts with animal species, and the historical changes in hunting and land tenure affecting the profile of animals currently encountered. We then report the results of clustering animals by contact frequency with human beings and evaluate the influence of gender, village, animal relative abundance, and human activity on contact frequency.

### Participant profiles

The profiles of participants in our data collection activities are detailed in Table 2.

**Table 2.** Participant profiles in data collection tools.

| Data collection tool | Gender |  | Age | Total |
| --- | --- | --- | --- | --- |
|  | F | M |  |  |
| Volunteer contact investigation | 7 | 17 | 18-30: 4<br>31-45: 8<br>46-59: 4<br>60+: 8 | 24 |
| Interviews | 25 | 63 | 18-30: 7<br>31-45: 32<br>46-59: 14<br>60+: 35 | 88 |

### Diversity of animal species, human activities and changes over time

#### Current human activities in forest and savanna environments

Our quantitative data yielded no differences in activity frequencies between the three villages, except for net fishing between villages B and C (Table 3). Agriculture and gathering were performed about half of the days, with significantly higher frequency performed by women. We did not observe differences in hunting frequency between the three villages.

**Table 3.**
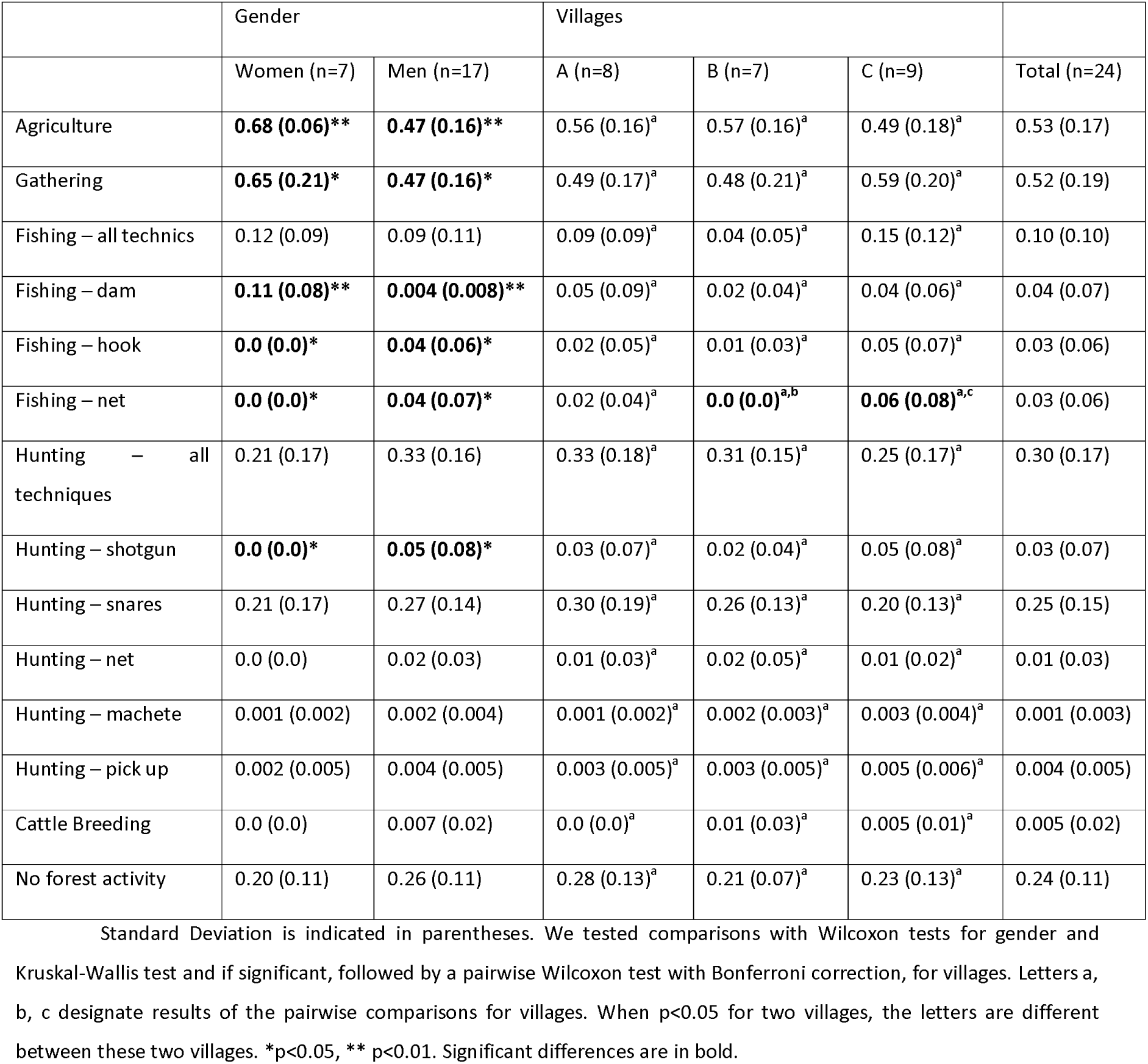
Mean frequencies of human activities by gender and village.

|  | Gender |  | Villages |  |  | Total (n=24) |
| --- | --- | --- | --- | --- | --- | --- |
|  | Women (n=7) | Men (n=17) | A (n=8) | B (n=7) | C (n=9) |  |
| Agriculture | **0.68 (0.06)\*\*** | **0.47 (0.16)\*\*** | 0.56 (0.16)<sup>a</sup> | 0.57 (0.16)<sup>a</sup> | 0.49 (0.18)<sup>a</sup> | 0.53 (0.17) |
| Gathering | **0.65 (0.21)\*** | **0.47 (0.16)\*** | 0.49 (0.17)<sup>a</sup> | 0.48 (0.21)<sup>a</sup> | 0.59 (0.20)<sup>a</sup> | 0.52 (0.19) |
| Fishing – all technics | 0.12 (0.09) | 0.09 (0.11) | 0.09 (0.09)<sup>a</sup> | 0.04 (0.05)<sup>a</sup> | 0.15 (0.12)<sup>a</sup> | 0.10 (0.10) |
| Fishing – dam | **0.11 (0.08)\*\*** | **0.004 (0.008)\*\*** | 0.05 (0.09)<sup>a</sup> | 0.02 (0.04)<sup>a</sup> | 0.04 (0.06)<sup>a</sup> | 0.04 (0.07) |
| Fishing – hook | **0.0 (0.0)\*** | **0.04 (0.06)\*** | 0.02 (0.05)<sup>a</sup> | 0.01 (0.03)<sup>a</sup> | 0.05 (0.07)<sup>a</sup> | 0.03 (0.06) |
| Fishing – net | **0.0 (0.0)\*** | **0.04 (0.07)\*** | 0.02 (0.04)<sup>a</sup> | **0.0 (0.0)<sup>a,b</sup>** | **0.06 (0.08)<sup>a,c</sup>** | 0.03 (0.06) |
| Hunting – all techniques | 0.21 (0.17) | 0.33 (0.16) | 0.33 (0.18)<sup>a</sup> | 0.31 (0.15)<sup>a</sup> | 0.25 (0.17)<sup>a</sup> | 0.30 (0.17) |
| Hunting – shotgun | **0.0 (0.0)\*** | **0.05 (0.08)\*** | 0.03 (0.07)<sup>a</sup> | 0.02 (0.04)<sup>a</sup> | 0.05 (0.08)<sup>a</sup> | 0.03 (0.07) |
| Hunting – snares | 0.21 (0.17) | 0.27 (0.14) | 0.30 (0.19)<sup>a</sup> | 0.26 (0.13)<sup>a</sup> | 0.20 (0.13)<sup>a</sup> | 0.25 (0.15) |
| Hunting – net | 0.0 (0.0) | 0.02 (0.03) | 0.01 (0.03)<sup>a</sup> | 0.02 (0.05)<sup>a</sup> | 0.01 (0.02)<sup>a</sup> | 0.01 (0.03) |
| Hunting – machete | 0.001 (0.002) | 0.002 (0.004) | 0.001 (0.002)<sup>a</sup> | 0.002 (0.003)<sup>a</sup> | 0.003 (0.004)<sup>a</sup> | 0.001 (0.003) |
| Hunting – pick up | 0.002 (0.005) | 0.004 (0.005) | 0.003 (0.005)<sup>a</sup> | 0.003 (0.005)<sup>a</sup> | 0.005 (0.006)<sup>a</sup> | 0.004 (0.005) |
| Cattle Breeding | 0.0 (0.0) | 0.007 (0.02) | 0.0 (0.0)<sup>a</sup> | 0.01 (0.03)<sup>a</sup> | 0.005 (0.01)<sup>a</sup> | 0.005 (0.02) |
| No forest activity | 0.20 (0.11) | 0.26 (0.11) | 0.28 (0.13)<sup>a</sup> | 0.21 (0.07)<sup>a</sup> | 0.23 (0.13)<sup>a</sup> | 0.24 (0.11) |

Standard Deviation is indicated in parentheses. We tested comparisons with Wilcoxon tests for gender and Kruskal-Wallis test and if significant, followed by a pairwise Wilcoxon test with Bonferroni correction, for villages. Letters a, b, c designate results of the pairwise comparisons for villages. When $p < 0.05$ for two villages, the letters are different between these two villages. \* $p < 0.05$ , \*\* $p < 0.01$ . Significant differences are in bold.

Most activities were conducted in the forest. Inhabitants practiced swidden (slash-and-burn) agriculture, producing primarily cassava (for subsistence and local trade), maize (for trade), plantain (subsistence and trade), and some fruits, legumes, and vegetables, such as pineapple and groundnuts. Inhabitants gathered edible plants (e.g. *Gnetum africanum*), mushrooms, caterpillars, medicinal plants, and building materials such as lianas for baskets or *Marantaceae* leaves for wrapping food.

Hunting was the third most performed activity, conducted with multiple techniques which targeted diverse animal species and entailed different exposures to biological fluids. The first, shotgun hunting, was practiced exclusively by men. This non-selective hunting technique targeted arboreal monkeys, duikers, antelopes, carnivores, and sometimes large rodents or birds. As one hunter acknowledged, “When you hunt with shotgun […] you target everything that crosses your path, even birds.” (140921-B)

Other techniques included snare trapping, net hunting, machete hunting and live collection. Trappers, both men and women, set up their snares in the forest and at the edges of their fields to capture medium-sized animals, such as large rodents, duikers, snakes, or carnivores (020419-C, 050819-A, 090921-B). Net hunting, an exclusively male activity, was performed only in the forest with the assistance of several dogs. This technique targeted duikers and large rodents (150921-B, 160921-B). Both women and men engaged in machete hunting, primarily for slow animals like pangolins and turtles (150921-B).

Both women and men also practiced fishing, although they employed different techniques in forest and savanna streams (091021-C, 150921-B). Women performed dam fishing (building a barrier in a running stream and emptying the water to capture fish), whereas men engaged in line and net fishing (Table 4).

**Table 4:** Mean number of contact events (Standard Deviation) – all contact types – and mean number of animal species (Standard Deviation) for all volunteers, by gender and village.

|  | Gender |  | Villages |  |  | Total<br>(n=24) |
| --- | --- | --- | --- | --- | --- | --- |
|  | Women<br>(n=7) | Men<br>(n=17) | A (n=8) | B (n=7) | C (n=9) |  |
| Contact events per volunteer over the study period (SD) | 3,979 (960) | 4,121 (1,156) | **4,737 (403)<sup>a</sup>** | 4,179 (1,192)<sup>a, b</sup> | **3,419 (1,113)<sup>b</sup>** | 4,080 (1,084) |
| Animal species (SD) | 33 (7.3) | 35 (5.0) | 31 (4.5)<sup>a</sup> | 37 (6.5)<sup>a</sup> | 35 (4.4)<sup>a</sup> | 34 (5.6) |
| Domestic animal species (SD) | 9 (0.4) | 9 (0.6) | 9 (0.5)<sup>a</sup> | 9 (0.8)<sup>a</sup> | 9 (0.3)<sup>a</sup> | 9 (0.5) |
| Animal species/day (SD) | 7.2 (1.8) | 7.6 (1.5) | **8.6 (0.9)<sup>a</sup>** | 7.4 (1.8)<sup>a, b</sup> | **6.5 (1.2)<sup>b</sup>** | 7.5 (1.6) |
| Domestic animal species/day (SD) | 4.5 (1.2) | 4.9 (1.1) | 5.8 (0.3)<sup>a</sup> | 4.6 (1.1)<sup>a, b</sup> | 4.1 (0.9)<sup>b</sup> | 4.8 (1.1) |

Only two of 24 volunteers raised cattle, doing so on ranches in savanna zones (040419-A, 020419-A).

#### Contacts with a large diversity of animal species

Our volunteers were in contact at least once with 61 different animal species, including nine domesticated species. These 61 animals were recorded in the following Orders: Artiodactyla (n=14 species), Carnivora (n=14), Rodentia (n=12), Primate (n=6), Chiroptera (n=3), Squamata (n=2), Afrosoricida (1), Anseriforme (n=1), Crocodilia (n=1), Columbiformes (n=1), Galliformes (n=1), Hyracoidea (n=1), Macroscelidea (n=1), Pholidota (n=1), Testudine (n=1), Tubulidentata (n=1). Over the five-month volunteer study, 97,917 contact events were recorded, with a mean of 4,080 contact events/volunteers (+/− SD=1,084; range=2299-6234). The volunteers reported contact with 7.5 different species per day on average (+/− SD=1.6, range=5.0-10.1), including 4.8 domestic species per day (+/− SD=1.1). We found no statistical difference between men’s and women’s contact events or species diversity per day, but we found that volunteers from Village A had a higher number of contact events and diverse animal species/day because of their higher contact frequency with domesticated species (Table 4).

#### Historical changes in hunting and wildlife profiles

Our qualitative interviews situated our results concerning current human contacts with diverse animal species in longer term processes. Elders recounted undomesticated faunal diversity has been subject to important changes over the past several decades, and that prior generations had more interactions with a higher diversity of animal species. As one elder man put it,

> “Past generations hunted more than today, with nets, guns, traps. [In previous times] the elders went 3 or 4 days into the forest to hunt with nets or traps. There were big hunting expeditions (*molao*) in the dry season. The last time we saw this here was in the 1990s” (090921-C).

Interview participants frequently characterized their current relations with forest and savanna animals (*nyama a mofuru* and *nyama a ntsio*, respectively) in terms of the past. They frequently complained about the declining numbers of undomesticated fauna over their lifetime, as evidenced by their current difficulties encountering and hunting specific animals (020419-C; 300319-B). Former hunters regaled us with past stories of returning from the hunt with 10 large rodents or duikers, an offtake that would be considered impressive today. Most elder male participants explained that current wildlife population depletion resulted from high hunting pressures during their youth or their parents’ lives (010419-B; 020419-C). Before the 1960s, they explained, obtaining firearms was easy, and selling meat earned quick money (260319-B; 120921-E), which decimated several monkey species (*Pilicolobus sp.*, *nkana* in Tio language; *Angola colobus, mvu; Lophocebus sp., nzela*) in the 1960s and 1970s. Subsequently, bongos (*Tragelaphus eurycerus, ngonoju*), panthers (*Panthera pardus, ngo*), black-fronted duikers (*Cephalophus nigrifrons, bokeli*) and giant pangolins (*Smutsia gigantea, nkau*) nearly disappeared, accelerated by easy accessibility of 12-gauge shotguns from the 1970s. As another retired hunter regretfully observed,

> “We have made bad use of the forest. Before, a hunter could kill 20 monkeys in one go. […] After 1972 there were a lot of hunters, a lot of shotguns, a lot of 12-gauges”. (120921-B)

#### Changes in land use and perceived consequences on faunal structure

For our participants, past land use dynamics also shaped their engagements with animals. Our historical interviews indicated that since the 1970s, several land tenures changes have affected ecological habitats and faunal structures. Just before DRC independence in 1960, cattle farming gradually expanded into savanna lands, which had until then been used for agricultural production and hunting. Expanding cattle raising thus appropriated these lands, displacing hunting and farming activities into forest lands. Hence, hunters conducted most activities in the forest, reducing their hunting of savanna-dwelling animals. In addition, the displacement of agriculture into forested areas increased pressures on available lands, leading to reduced fallows, from 5-10 years to 2-3 years. Not only have decreased fallows led to decreased productivity of agricultural lands, but they also produced what some informants called “dead forest”, that is, the colonization of fallowed lands by the invasive *Chromolaena odorata* (*felele*) and ferns, primarily *Pteridium spp.* (*enye*). As focus group participants explained,

> – “After we work the fields for several years, ferns (*enye*) emerge.”
>
> – “Yes, first we have the forest (*mokuna*), which is transformed into the fallow (*evuu*), which brings the “flower” (*felele*), after *felele* comes the ferns: and there it’s the end!” (140921-B, focus group).

Our participants explained that fallowed lands converted more easily into savanna, thus affecting animal diversity and abundance. They suggested that fern colonization of these sites could create habitats favorable to large rodents –– cane rats (*Thryonomys swinderianus, nsili*), Emin’s pouched rats (*Cricetomys emini, nkuli*), brush-tailed porcupines (*Atherurus africanus, nkeon*), and small carnivores (140921a-B).

### Types of contacts and categorization of animal species

#### Categorization of animal species according to their contact profile

We categorized animal species according to their contact profile. Mean frequencies for contact with each species and each type of contact are detailed in Supplementary Table 2. The first two axes of the PCA explained 81% of the variance. Visualization of the 14 types of contact within the two first axes shows high correlation between two broad contact types (Fig 2): all physical contacts plus “seen dead”, and all environmental (direct and indirect, except for “seen dead”) contacts. We therefore consolidated these diverse contacts into two categories –– physical and environmental. We reassigned the first two coordinates of the PCA to each animal species and performed HCAs. We obtained three clusters of animal species according to their contact profiles (Fig 3 and Supplementary Table 3). The pairwise comparisons of these clusters indicated significant differences (Fig 4 and Supplementary Table 4 and 5). The first cluster (n=46 species) represented most animal species and was characterized by low contact frequency for all types of contact. The second cluster (n=6) was characterized by animals with which humans had the highest physical contact rates; the third cluster (n=9) had the highest frequency of environmental contacts. This latter cluster is composed of all domesticated species (except rarely-raised pigs, *Sus scrofa domesticus*, *ngulu*) and a commensal rodent species (*Mastomys natalensis*, *mpu*).

**Fig 2.**
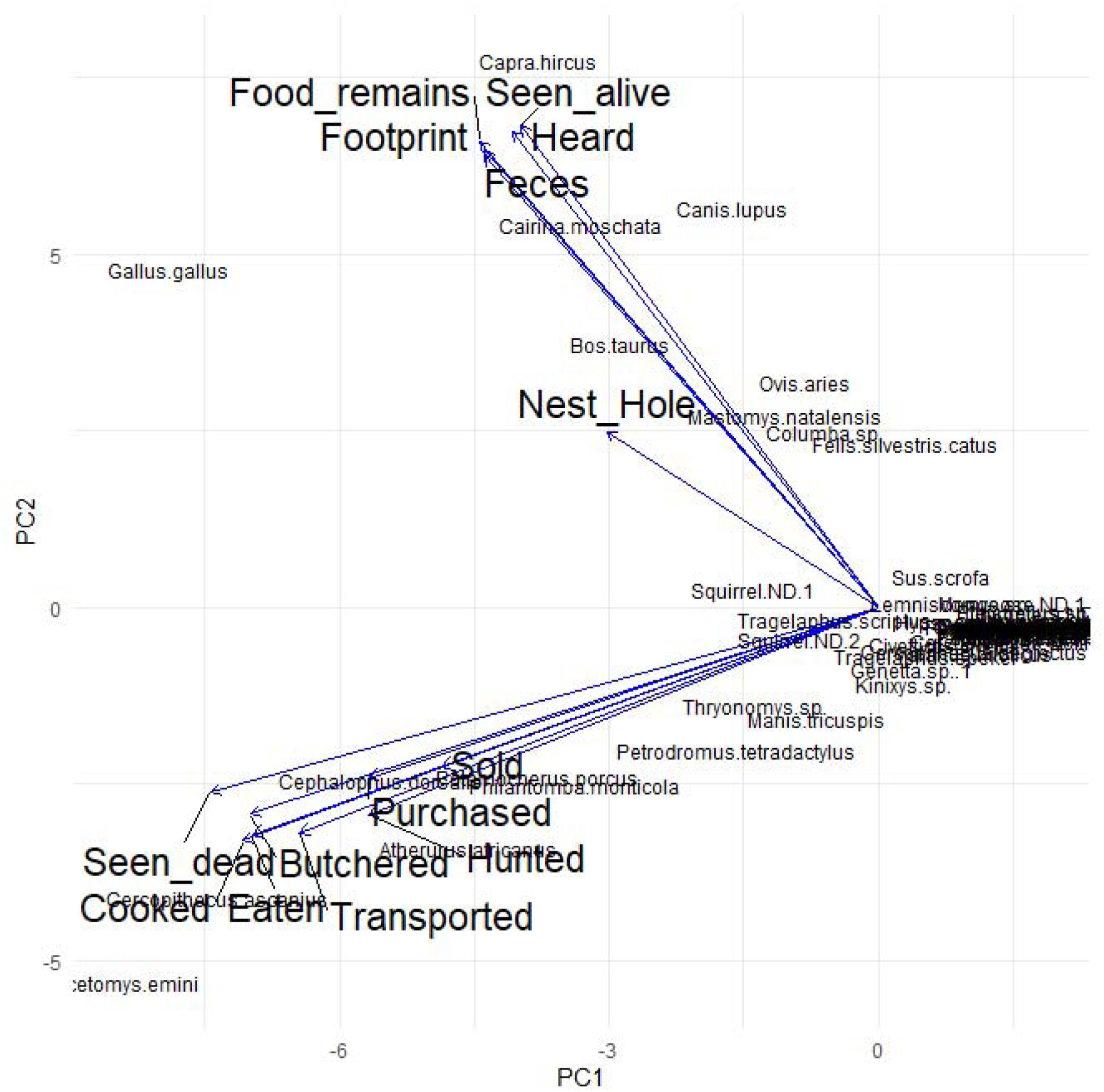
Principal Component Analysis of animal species according to frequency of contacts with humans for each type of contact. The two first axes explained 81% of variance.

**Fig 3.**
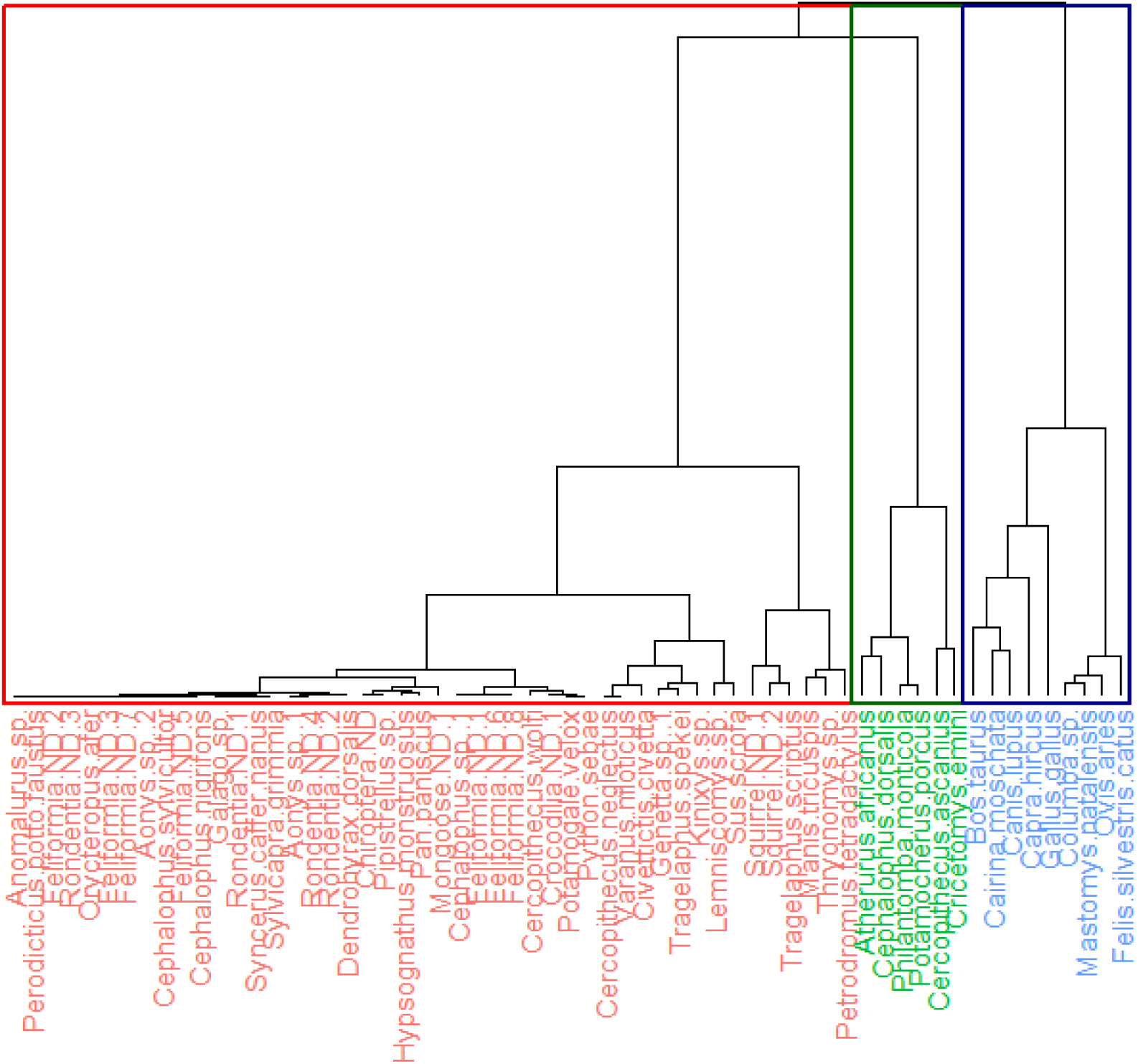
Hierarchical cluster analysis of animal species according to projected coordinates of the two first axes of the PCA.

**Fig 4.**
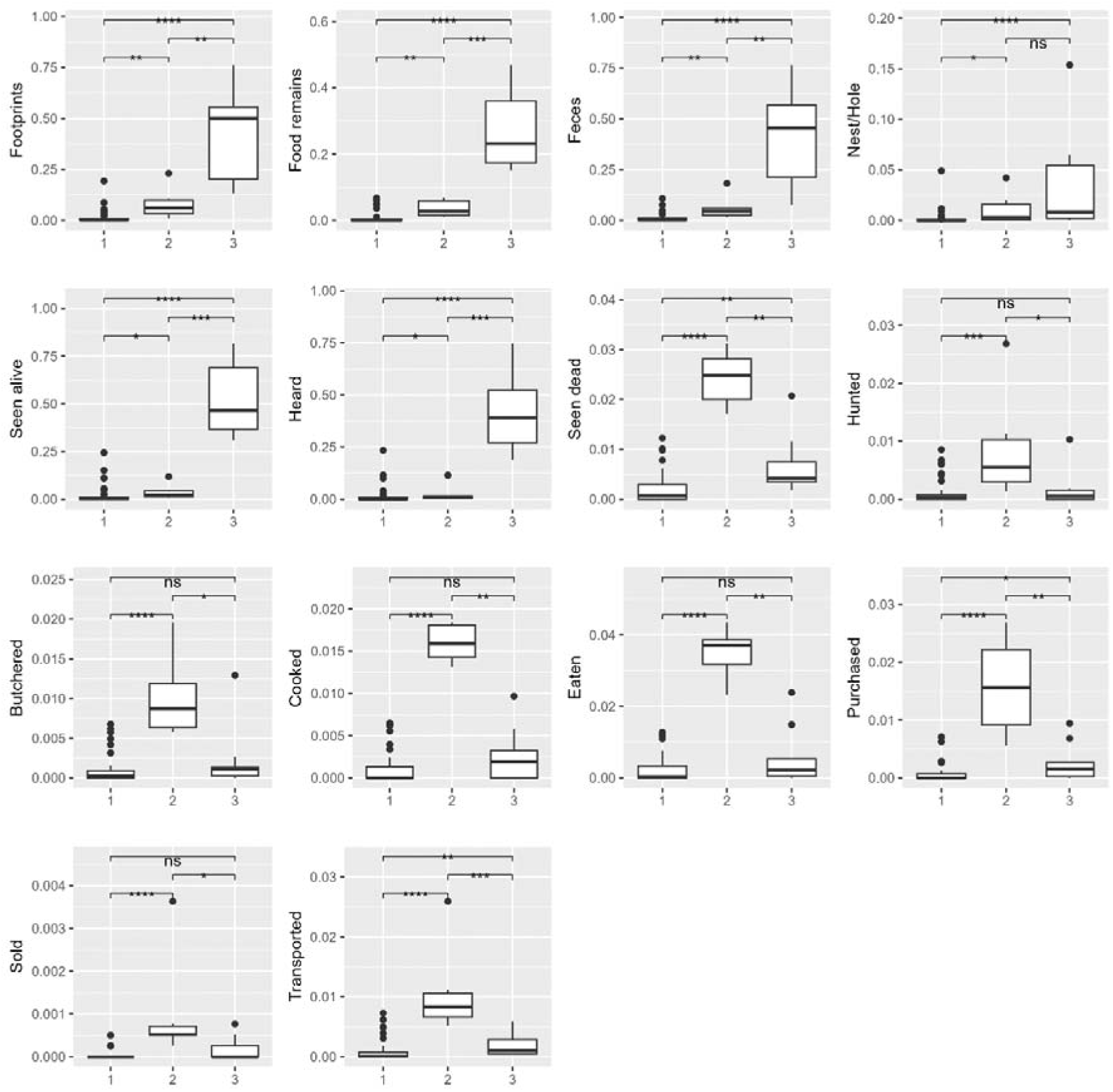
Boxplot comparisons of contact frequencies for each type of contact among the three clusters obtained with the HCA. For each boxplot, the black bar represents the median; white boxes display the interquartile range (25-75th percentile). The lower limits of the vertical lines represent the smallest value within 1.5 times the interquartile range below the 25th percentile, and the upper limits represent the largest value within 1.5 times the interquartile range above the 75th percentile. Black dots are values outside of these limits. ns=not significant; * p<0.05; ** p<0.01; ***p<0.001; ****p<0.0001.

#### A web of environmental contacts for cluster 3 animals

Cluster 3 animals, largely domesticated species, lived near or with human inhabitants of the three village sites. We observed domestic animals roaming freely in these villages, sometimes ranging as far as the forest or savanna edges abutting on these villages. These domestic animals had different ranging behaviors and were treated differently by their owners.

Chickens and ducks remained close to households to which they belonged, and they sometimes entered homes in search of stored maize. At sundown, to protect flocks from theft or mongoose attacks, women owners coaxed their poultry into enclosures next to their houses or into separate kitchen structures. Our participants explained that poultry required little time investment, management, or physical contact. Current flock sizes, ranging from 2 to 10 birds, were much smaller than those of 20 or 30 years ago because of recurrent epidemics and owners more limited purchasing capacities (010821-A).

Goats, and to a lesser extent sheep, were also free ranging. Goats and sheep herds tended to mingle with those of different owners during the daytime. They also crossed paths with village poultry. At times, they entered open houses in search of drying cassava. Human contact with goat and sheep feces occurred frequently, and children regularly cleaned this excrement and discarded it in open waste areas next to household concessions.

Pigs were rarely raised, and thus environmental contact occurred rarely. As several participants explained, village inhabitants ‘“evicted” them because they host parasites and damage property’ (060819-A; 091721-B).

Many people considered certain domesticated animals (e.g. sheep, goats, and ducks) to be *ngeyi*, an expression meaning “disgusting” or “repulsive” and evoking filth and impurity. Hence, village inhabitants refrained from touching or consuming these animals, although older people tended to express this disgust and avoidance more frequently than younger inhabitants.

Additionally, domestic animals (*nyama a bola*, litt. “animal of village”) raised for meat generally functioned as a “living bank accounts” and were thus rarely eaten (and hence touched). Cattle were frequently sold and exported for revenue except during local festivities. An owner could slaughter a goat to obtain urgently needed cash (for health expenses, funerals), selling small portions of the meat locally. Inhabitants sold their sheep to Muslim merchants from neighboring Republic of Congo, notably in Village A, closest to the market town and the Congo river (040819-A).

Human-dog engagement with dogs differed from that with other domesticated animals. Village inhabitants used dogs for solitary spear or collective net hunting and to guard their homes. During the day, dogs roamed throughout the village searching for other dogs and leftovers in waste areas (the same sites where animal wastes were discarded and young children defecated (210921-A, 110921-C)). Dog owners engaged in physical contact with dogs primarily when hunting and when managing, caring for, and feeding them. Dogs also maintained close environmental contact with their owners, sleeping in the main house or kitchen area.

Very few cats were kept in villages A, B, or C. The few cats observed moved between houses in search of mice.

The sole non-domesticated species in cluster 3 was *Mastomys natalensis* (mouse). Our three-village survey with all households found that all respondents except three had mice in their houses, but that housing (roof and wall) materials had no effect on mouse presence. Our volunteers also reported important environmental contact frequencies with *M. natalensis* (notably, “seen alive” mean=0.31, SD=0.20, min=0, max=0.68). Village inhabitants in all sites considered household mice to be a pest *par excellence*, partly because of their destructiveness, but also because of their perceived impurity, in contrast to forest mice and rats. Survey and interview results showed that people did not eat house mice, although forest and savannah mice provided frequent supplemental meals for children (090821-C). Village inhabitants sought to control *M. natalensis* with human anti-inflammatory drugs, rat poison, salt, rarely cats, or by bludgeoning them with sticks or branches (150921-B).

Our participants actively avoided touching living or dead house mice or their feces. They would pick up a mouse carcass using a piece of plastic or cloth and then discard it in a refuse dump at their concession edge. As two participants noted,

“The mouse is considered the enemy of humans, this is why no one accepts it in their house” (140921b-B)

“It’s an animal of bad faith. It does not come to eat, but to destroy” (080921a-C).

This avoidance of physical contact resulted, participants contended, because mice frequented latrines and could transmit illness. These illnesses went unspecified, although infrequently participants referred to typhoid or fever (150921-B). Some used mosquito nets around their beds to avoid contact with mice or their feces. They claimed that mice were wily, blowing or nibbling on the feet of sleeping humans to divert attentions away from other mice destroying human possessions.

#### High physical contact with cluster 2 animals

Our participants had the highest physical contact frequencies with cluster 2 animal species, which included multiple forest animals: two rodent species (*Atherurus africanus* and *Cricetomys emini*); three Artiodactyle (even-toed) species (*Philantomba monticola, Cephalophus dorsalis, Potamocherus porcus*) and one primate species (*Cercopithecus ascanius*). Our interviewees reported that these species were commonly hunted and killed. Human exposure to a hunted animal’s bodily fluids depended on the hunting technique used and game size (Table 6).

**Table 6:** Types of contacts according to species of the cluster 2 (high physical contacts). *[M]: men; [F]: women.

| Species with high physical contact | Main situation of contact during capture [by whom]* | Contact through slaughtering and transport | Butchering | Meat sharing and sales (with implications for the scale of physical contact) [by whom] |
| --- | --- | --- | --- | --- |
| **Brush-tailed porcupine**<br>(*Atherurus africanus*) | Snare trapping [M,F] | Killed by snare or slaughtered by machete, and transported whole to village [M,F] | Butchered in village [M] | Limited sharing (household level) |
| **Emin's pouched rat**<br>(*Cricetomys Emini*) | Fire or flushing out, with or without dogs [M] | Killed by human or dog, transported whole to village [M] |  |  |
| **Blue duiker**<br>(*Philantomba monticola*) | Collective net hunting with dogs [M] | Killed by dog or human (piece of wood or machete), handled fresh [M] | Butchered at hunting site [M], with fresh butchered pieces transported to village [M] | Extensive sharing (hunting team) on the hunting site |
|  | Snare trapping [M,F] | Killed by snare or slaughtered by machete, and transported whole to village [M,F] | Butchered in village [M] | Limited sharing (household/kin group level) or sold in pieces |
|  | Shotgun [M] | Killed and transported whole to village [M] | Butchered in village [M] | Limited sharing (household/kin group level) or sold in pieces |
| **Bay duiker**<br>(*Cephalophus dorsalis*) | Shotgun [M] | Killed, with quarters transported to village [M] | Butchered in village [M] | Extensive sharing or selling (kin group and village level) |
|  | Collective net hunting with dogs [M] | Killed by dog or human (machete), handled fresh [M] | Butchered at hunting site [M], with freshly butchered pieces transported to village [M] | Extensive sharing (hunting team) on the hunting site |
| **Red river hog** (*Potamocheirus porcus*) | Shotgun [M] | Killed, with freshly butchered pieces transported to village [M] | Butchered at hunting site [M] | Extensive sharing and selling (kin group and village level) |
| **Red-tailed monkey** | Shotgun [M] | Killed and transported whole (fresh) to village [M] | Butchered in village [M] | Limited sharing (household level) |
| (Cercopithecus Ascanius) |  |  |  | or sold in pieces |

Generally, hunters avoided drawing blood during the hunt. If a hunting dog did not kill the game directly during net hunting, the hunter would break the neck of small animals (e.g. blue duiker) and kill larger game with a heavy blow to the animal’s head to prevent blood spillage during (150921-B).

Our participant-observations showed complex meat sharing practices from individual hunting and trapping, and thus uneven exposures to game meat and bodily fluids. The gift of wild meat, reinforcing social cohesion with kin, friends, and local authorities, followed specific butchering rules and “rights” that defined what animal parts or organs a hunter owed to specific recipients. How undomesticated animal meat was shared depended on the hunting techniques used and species killed, so that sharing obligations for small animals was less strict than for larger ones. After a successful hunt or trap visit, a hunter would bring game home to be butchered discreetly, by himself or one of his sons, but never by household women. The hunter sent fresh meat parts or organs to specific relatives, so that a hunter killing a monkey with a shotgun would send its heart to his mother, kidneys to his father, its chest to his maternal uncle, its neck to his brothers. The hunter and his household would consume the rest of the animal (090921-C). In general, larger wild meat led to a more extensive sharing and selling of smaller pieces among a larger group, while small animals were consumed among immediate family members. Moreover, in response to meat scarcity, small animals tended to be hidden from the community to avoid jealousy and demands to share.

Local inhabitants finding animals visibly suffering from illness would kill and eat them; they might also consume game found dead in the forest, unless the carcass was too decomposed (190921-A).

Finally, collective hunting entailed animal bodily fluid exposure for all participants. Before returning home, hunters butchered all game killed at the forest edge (a practice called *ozamba*), then grilled and ate some meat before sharing the rest among themselves according to precise rules. Following their return home after the hunt, the hunters would field meat demands from families and neighbors. They would hand over meat that they had kept to their wives or sometimes kin elders to be cooked.

#### Infrequent contact frequency with cluster 1 animals

Cluster 1, which included some 46 animal species, was characterized by lower environmental and physical contact frequencies in comparison to the other clusters. Our relative abundance calculations for 17 species in this cluster revealed a significantly lower relative abundance compared to cluster 2 species (Mann-Whitney test, p<0.05) and a tendency with cluster 3 species (Mann-Whitney test, p=0.068). Certain cluster 1 species, however, had higher relative abundance compared to certain cluster 2 and 3 species but much lower contact frequencies.

Analyses of our qualitative evidence provided additional insight into low contact frequencies with cluster 1 species, most notably a gendered avoidance of certain animals because of shared social norms or because of individual or family sensitivities, past experiences, or disgust. For example, women have long avoided carnivore species (10 cluster 1 species) because of their smell (190921-A). Two carnivore species cluster 1, *Civettictis civetta* or Herpestidae gen. sp. had high relative abundance, although men and women volunteers did not display significant differences in contact with them. Women, however, consumed these species half as frequently as men.

Inhabitants of the three villages avoided certain species for other reasons. Pregnant women, for instance, steered clear of monkeys so that their newborns would not resemble one (190921-L). Adults typically avoided bonobos (*Pan paniscus*; ebubu), hyrax (*Dendrohyrax dorsalis*; ebuya), and Potto (*Perodicticus potto*; ekaru) because these animals lacked a tail, which participants contended rendered them more like humans. Local myths accorded bonobos a special place, insisting that they had human forms in ancient times (020419-C; 290319-A; 300319-B). As one participant recounted,

“The ban on eating the bonobo has been in place since time immemorial, because the bonobo has characteristics resembling those of humans. It is said that the bonobo was originally a human who fled into the forest and avoided living with people in the village. And all animals have tails except the bonobo. Its hands look like a human’s. It also has a bad smell…” (290319-A)

More broadly, during interviews village inhabitants labeled about 20 animal species as *ngeyi* (*disgusting*, *repulsive)*. Among these species, 6 appeared in cluster 3 (five domesticated animals, and *Mastomys natalensis*, mouse present in houses), and the other 14 appeared in cluster 1 (Fig 5). They include four carnivores (*Nandinia binotata, Civettictis civetta, Genetta sp., Herpestes sp.*), three primates (*Pan paniscus,* Perodicticus *potto*, *Galago sp.*), but also *Lemniscomys sp., Petrodromus tetradactylus, Aonyx capensis, Varanus niloticus, Dendrohyrax dorsalis, and Pipistrella sp*. Certain rodents (*Lemniscomys sp., Petrodromus tetradactylus*), although considered dirty by adults, were nonetheless often hunted and eaten by children.

**Fig 5.**
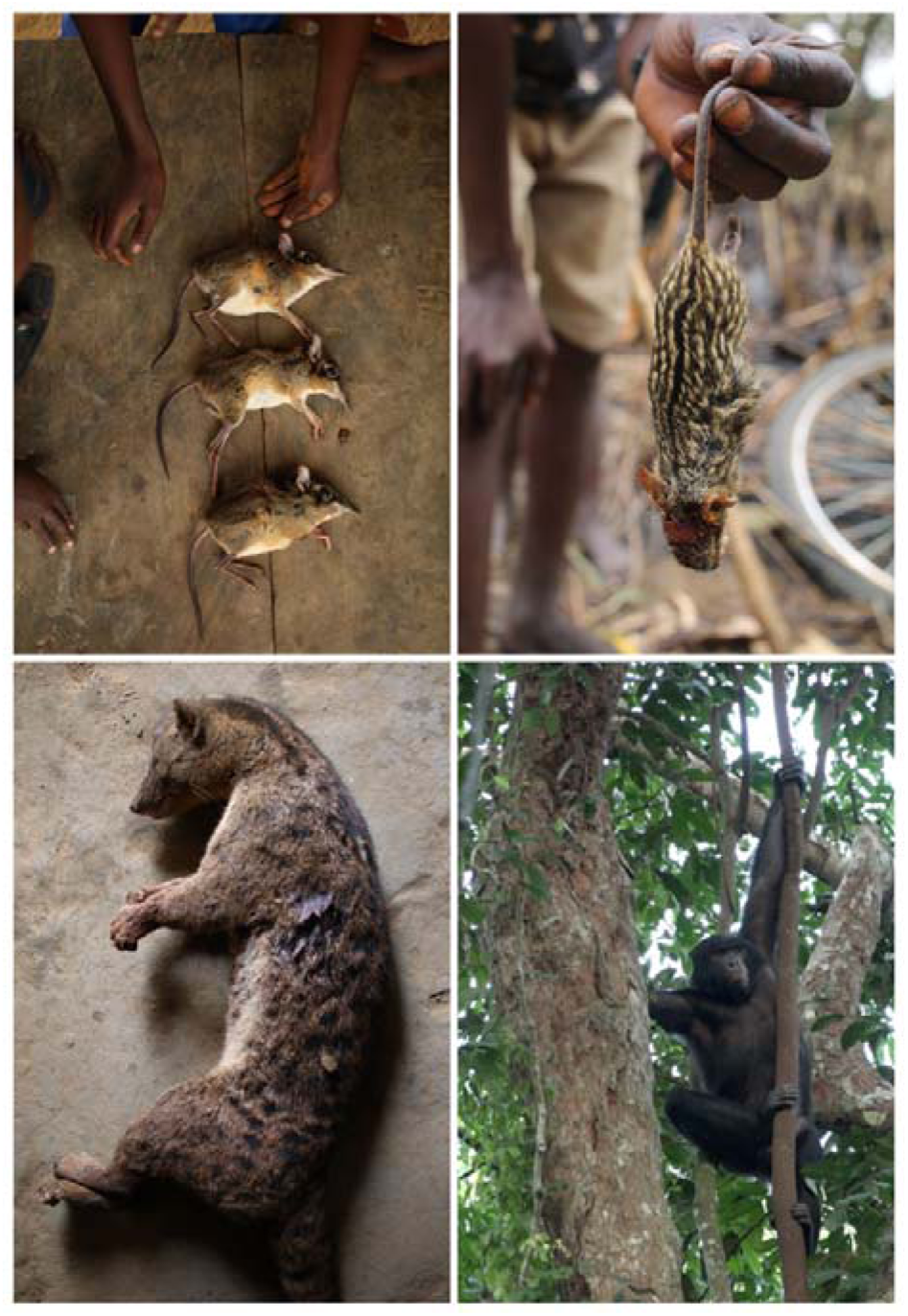
Cluster 1 animals (top left: Petrodromus tetradactylus; top right: Lemniscomys sp.; bottom left: Nandinia binotata; bottom right: Pan paniscus) (credit: Author 1)

#### Influence of village, gender, species relative abundance, and human activities on contact frequencies

To test the influence of village, gender, species abundance, and human environmental practices on contact frequency with animals, we performed six GLMMs, one for each interaction between the contact category from the PCA (physical or environmental contact) and the animal species cluster from the HCA (cluster 1 with low contact frequency; cluster 2 with high physical contact frequency; and cluster 3 with high environmental contact frequency). We obtained relative abundance values for 32 of the 61 animal species recorded in the contact dataset and thus performed these GLMMs on these 32 animal species. Table 7 synthesizes the results (see Supplementary Information 2 for detailed result).

**Table 7:** Synthetic results of GLMMs.

|  | Cluster 1 |  | Cluster 2 |  | Cluster3 |  |
| --- | --- | --- | --- | --- | --- | --- |
|  | Environmental contacts | Physical contacts | Environmental contacts | Physical contacts | Environmental contacts | Physical contacts |
| Village | Village B weak positive | Village A, weak negative | / |  | Village A, positive, Village B tendency, positive | / |
| Gender |  |  | / |  |  | / |
| Relative abundance | weak positive | weak positive | / | weak positive | weak positive | / |
| Hunting – all techniques |  |  | / | weak positive |  | / |
| Fishing – all techniques |  | tendency, weak positive | / | tendency, weak positive |  | / |
| Gathering |  |  | / |  |  | / |
| Agriculture |  |  | / | negative |  | / |
| No forest activity | negative |  | / |  |  | / |

When full model was significantly different from the null model, we report the potential effect of each variable. A blank cell means no significant effect. We used “/” to indicate that the full model was not significant compared to the null model.

Within the six GLMMs performed, four models were significant in comparison with the null model. Among these four models, gender had no influence on contact frequency. Village site did influence contact frequency but displayed no clear pattern. Relative abundance had a significant and positive association with contact frequency (weak effect) for all the four models. Hence, more abundant species located closer to villages displayed higher physical and environmental contact for cluster 1 species, higher physical contact for cluster 2 species and higher environmental contact for cluster 3 species, with human inhabitants.

Cluster 2 species had higher contact frequency with humans heavily engaged in hunting and fishing, and lower contact frequency with those more heavily engaged in agricultural activities.

We thus identify several factors and processes associated with diverse human-animal contacts in an ecologically varied zone and its categorization of contact profiles.

## Discussion and conclusions

Overall, our participatory activity and contact survey revealed that across the three villages, participants were in physical and environmental contact with 61 different animal species. We also categorized animal species according to their contact profile with humans, without preconceived categorization. Our qualitative evidence situated these contacts in longer-term historical changes related to hunting and land use. Finally, our modeling analysis of gender, village, relative abundance and human activities on animals clustered by contact frequency yielded few predictors of contact frequencies. The following addresses each of these findings in engagement with broader literatures on human-animal contacts and their implications for zoonotic pathogen exposure.

### Human activities

We initially evaluated whether activity frequencies of participants differed by village and gender. Here we found no differences in activity frequencies between the three villages. Women performed agriculture and gathering more frequently than men across the three villages. Although these forest edge societies do display an important gender division of labor, we found that women also contributed to hunting activities, frequently visiting snare traps set by their husbands near fields and bringing home small trapped mammals.

Our findings largely convene with the range of human activities in forest societies in central Africa [40–43], although we expect that the proportions of labor expended for agriculture, hunting, gathering, and other activities would vary across societies. Nonetheless, whereas some studies of human activities in the forest have oversimplified the range of practices, we show that our participants performed wide-ranging activities in the forest. Hunting practices, from hunting with dogs to shotgun hunting, reflect practices elsewhere in central Africa. In addition, although women’s hunting practices have frequently been overlooked in hunting literatures in Africa, multiple studies have reported or highlighted such activities among women [44–46].

### Diversity of contact with animal species

We found that our participants were in physical and environmental contact with a very high diversity of animal species (61), with a higher number of contact events and diverse species per day in Village A than in the other villages. This distinctive feature of Village A animal contacts resulted from higher contact frequencies with domesticated animal species in a more fragmented forest, and where small-scale sheep and goat production was more developed because of proximity to the riverine market town.

Here our results depart from and convene to a broader literature on human-animal contact. Older contact studies have tended to evaluate human contacts with selected species or animal host reservoirs of specific pathogens, thus limiting the diversity of animal species documented [14,31,47–49]. More recent contributions, however, have expanded analyses of human-animal contacts to account for a range of domesticated and undomesticated animals [16,50,51]. The high diversity that we identified may differ from prior studies for multiple reasons: some studies develop data collection tools that request participants to report only their contacts with selected animal species; others aggregate animal categories (e.g. “monkeys”, rather than specific nonhuman primate species); still others may have been conducted in sites with less animal diversity.

### Clustering of animal species by contact

Rather than imposing pre-conceived categories on our analyses of human-animal contact frequencies, we categorized each animal species with which our participants had contact by their contact profile. Our PCA analysis yielded clustering into three distinct groups: cluster 3 animals, with a high frequency of environmental contacts (including the commensal rodent species *Mastomys natalensis* and all domesticated species except for pigs, which are rare in this region); cluster 2 animals, with high physical contact rates (which included a large rodent and small mammals); and cluster 1 animals, a wide range of undomesticated mammals with which our participants had infrequent physical or environmental contact.

This clustering constitutes a departure from human-animal contact literature, which has targeted selected animal species or pre-determined aggregated categories of animals [16,50,51]. Elsewhere, we have argued for abandoning analyses based on pre-conceived categories of “wild” and “captive” animals, which are neither homogeneous nor stable across diverse settings [52]. Here we would extend our reflections, arguing that we should use category labels critically and consider how grounded experiences can shed light on alternative categorizations. Although our cluster 3 species were roughly comprised of “domesticated” and commensal rodent species and cluster 2 species were frequently hunted mammals, these clusters emerged from nearly real-time human contacts, evaluated by frequency and type. The highest risk of transmission would, we anticipate, occur between humans and clusters 3 and 2 animals based on prior studies examining potential risks resulting from human-animal contact [16,53,54].

These clusters thus reveal critical insight into the risks incurred over time through the types and frequencies of contact with domesticated, commensal, and undomesticated animals. Although this manuscript does not report on the pathogens carried by animal species in the three village sites and transmitted to human inhabitants, we show the real, cumulative risks of zoonotic spillover posed by animal clusters over time. The risks of pathogenic transmission associated with cluster 1 animals, for instance, were minimal because our participants rarely had physical or spatial contact with these species. By contrast, risks of pathogenic transmission through physical contact substantially increased with cluster 2 species because our participants hunted, butchered, and prepared them most frequently. Participants would incur elevated cumulative risk for environmentally persistent pathogens with cluster 3 species because of their frequent spatial contact with such species. These clusters, then, offer potential guidance for surveillance systems, to monitor only those species with which humans have most frequent physical or environmental contact. More broadly, in documenting contact types and frequencies in near real time with animal species, rather than focusing on a single group of animals (e.g. rodents or nonhuman primates) or on a pre-selected list of animals, our study provides one effective means of evaluating real, cumulative risk of zoonotic transmission in a fragmented forest zone.

### Factors influencing contacts and potential risk of zoonotic pathogen exposure

Among the variables tested in our multivariate model, we have relatively little that allows us to explain contact frequency. We found no effect of gender on contact frequency in the significant models, and no clear pattern for an effect of the village.

Frequencies of human activities were associated with physical contact frequencies with cluster 2 species. We found no differences in activity frequencies between our three villages. Our finding concerning gender on contact frequency may result from the fact that women performed agriculture and gathering most frequently across the three villages and contributed significantly to hunting activities. Forest edge societies do display an important gender division of labor, but women’s participation in hunting (by checking traps) echoes findings of studies elsewhere in central and west Africa [43–46,55]

We also found that relative abundance of animal species is associated with contact frequency: those species in higher abundance and closer to villages have higher contact frequency with humans. This result converges with a prior study on contact with nonhuman primates in Cameroon [15]. This effect, however, is weak and cannot fully explain the factors shaping human-animal contact. The results of our multivariate model illustrate that human-animal contact frequency is highly complex and difficult to distill into a discrete set of independent variables.

### Historical changes and contemporary understandings shaping human-animal contacts

Our ethnohistorical and ethnographic analyses suggest that longer-term historical processes and contemporary understandings of specific animals shaped human-animal contact, and more broadly, the potential risks of exposures to zoonotic pathogens.

Changing animal raising practices and land use appeared to frame patterns of human contact with diverse animal species. Allowing domesticated animals to range freely is widely practiced throughout the African continent, and thus it is not surprising that cluster 3 composition reflected frequent environmental contacts [56]. The advent of large– and small-scale cattle farming from the mid-twentieth century appears not only to have shape these contacts, but generated other alterations in land use, ecological zones, and human-animal contact. Our oral histories revealed that cattle farming precipitated substantial land use changes, transforming savannas into grazing lands for cattle, and leading to significant dispossession of arable savanna lands among women and the displacement of women’s cultivation and hunting activities from savannas into forest sites. These land use changes, our participants recounted, affected forest farming practices (shorter fallows) and additional ecological transformations, notably “dead forests” with different botanical composition and populated with certain rodent species, (brush-tailed porcupine, Emin’s pouched rat) with which our participants had considerable physical contact. Hence, the introduction of cattle raising had long-term ecological consequences, which in turn affected human-animal contact and potential increased risks in zoonotic transmission. Our analysis reveals, then, that hunting is not the only factor shaping human-animal contact, proposing that changes in land use and animal husbandry can reshape the frequencies and types of contacts to specific animals.

Nevertheless, hunting pressures may have contributed to frequent human contact with these large rodents. Our older participants contended that overhunting depleted large, undomesticated animals, the consequence of the arrival and democratization of hunting rifles, as observed elsewhere [57]. This reported disappearance of larger animal species may explain why our participants had infrequent contact with wide ranging animal species (cluster 1) and frequent physical contact with small mammals (cluster 2). In Africa and elsewhere in the world, land use and animal husbandry changes, as well as long-term hunting intensification have been shown to transform the profiles of animal populations [58]. The replacement of large mammals by smaller ones echoes insights from other studies; large scale investigations show that hunting pressures and land use changes reduce nonhuman mammal diversity, facilitating increasing populations of robust, rapidly reproducing small mammals, including rodents [59–63].

In addition, human-animal contacts also seem to be shaped by human perceptions and uses of these animals. Our participants explained that they actively avoided certain animal species, some of which were understood to be “unclean”. These avoidance practices could explain the lack of physical contact with domestic animals and commensal rodents, as well as highly infrequent contact with certain cluster 1 animals. Several studies elsewhere in central Africa have explored the sociocultural understandings affecting contact with animals [64–67].

Overall, this multidisciplinary investigation yields broad, substantive insight into the diversity of human contacts with animals in this fragmented forest zone. We examined human contacts with many animal species, and through our analyses, indirectly shed light on cumulative risk of exposure to zoonotic pathogens. Zoonotic spillover risks are not equally distributed across all animal species. Although we do not evaluate direct spillover risks, we showed that human inhabitants of our study zone have frequent contact with a relatively small group of animals, both wild and domesticated. For this reason, surveillance of animal species in clusters 3 and 2 would be a crucial measure in anticipating zoonotic spillovers and in curbing outbreaks at their source. Human activity and relative abundance of animal species in proximity to villages are associated with contact frequencies, but the associations are not sufficiently strong to capture these complex dynamics.

Finally, our study sheds light on the complex historical processes and contemporary conditions that frame human-animal contact. “Contact” is not a singular, discrete event, but rather shaped by larger socioecological processes and sociocultural perceptions [12]. Our ethnohistorical and ethnographic approach generates new insight here, capturing these complex dynamics. Such an approach should be integrated more consistently into future studies.

### Limitations of the study

This study has several limitations that necessitate some caution in interpretating our results. First, our participatory contact survey had some important shortcomings. The sample size of volunteers was relatively small and heavily biased in favor of literate men. It cannot be considered representative of the three village populations nor of the province, even though our limited sample generated a considerable quantity of data. Future studies could employ less labor-intensive tools to cover a representative sample of village and provincial populations.

Our contact study covered only five months of the year, and thus our activity and animal contact data may be biased. Nevertheless, inhabitants of these village sites conduct farming, hunting, and fishing year-round, even if specific techniques associated with these activities changed across the seasons. Moreover, animal species of this area can follow some seasonal patterns but with no major changes of their distribution area and habitat use.

Although our participatory contact survey generated highly granular, near real-time reporting of contacts, it depended on participants’ individual ecological knowledge, as well as their attention to and recollection of animals and animal traces encountered. We sought to compensate for inattention and recall problems by careful training of volunteers and frequent visits from a field supervisor to ensure robust field data. Finally, this tool relies on a highly constrained definition of “contact”, that is physical or spatial. Local cultural understandings of contact were not integrated into the tool. Forest inhabitants, for instance, frequently use their sense of smell to detect or identify animals and plants, and they consider odor to be a form of interspecies contact. Nevertheless, we concluded that integrating other understandings of contact would render our participatory collection even more complex than it already was and risked confusing participants and rendering our results less reliable.

Furthermore, certain key animal species were missing from our transect survey. Heavy workloads and limited resources prevented our field team from evaluating numerous bat or monkey species. Future investigation should include these species.

Finally, this article addresses only risks associated with contact, not actual exposure to and infection with zoonotic pathogens. Such epidemiological and serological analysis is beyond the scope of this study.

## Data Availability

All data produced in the present work are contained in the manuscript, except for the qualitative data because ethics approvals do not permit the sharing of this data because it could compromise the confidentiality of research participants.

## Acknowledgements

We sincerely thank the 30 volunteers and other participants who contributed their time and reflections to the study. We are also grateful to the customary chiefs of the three study sites for their authorization and support for our field research. We thank researchers and personnel at INRB and Mbou-Mon-Tour for their field assistance and support. Muzungu Ngofuna and Emmanuel Sando Mbobi who provided essential research assistance in the field. We also thank Flora Pennec for the forest cover and edge density analysis.

Finally, we thank the Inception Program and the CIFAR Humans and the Microbiome program for funding this study. Inception Program funds (grant No. PIA/ANR-16-CONV-0005) are provided by the French Agence Nationale de Recherche/Investissements d’Avenir.

## Notes

### Competing Interest Statement

The authors have declared no competing interest.

### Funding Statement

This study was funded by the Inception Program and the CIFAR Humans and the Microbiome program. Inception Program funds Grant PIAANR16CONV0005 were provided by the French National Research Agency, Investments in the Future

### Author Declarations

The Institut Pasteur Institutional Review Board Decision No. IRB2018.08 and the National Committee of Health Ethics for Health of the DRC Decision no. ESP.CE.2019 and ESP.CE.045/2021 gave ethical approval for this work.

